# Lens Thickness and Its Association with General and Ocular Parameters in Healthy Subjects

**DOI:** 10.1101/2022.04.11.22273740

**Authors:** Yi Cao, Fucheng Liao, Yushen He, Yujuan Huang, Lijun Zhou, Xiangbin Kong

## Abstract

The crystalline lens lens thickness playing an essential role in maintaining normal visual function, but little attention is paid to the lens thickness. The purpose of this study is to document the normative values of lens thickness (LT) in healthy subjects of Southern China and to evaluate its associated factors. This was a prospective, clinic-based, observational, cross-sectional study. A total of 526 eyes from 263 healthy subjects aged between 5-84 years (mean age, 38.48 ± 22.04 years) were included in the study. All subjects underwent detailed ophthalmic examination, including the measurement of ocular biometric parameters by LenStar LS900. The study focused on LT and its association with general and ocular factors. The generalized estimation equation (GEE) model was used for statistical analysis. The Mean LT was 4.01 ± 0.57 mm(95% CI, 3.97 - 4.06; median, 3.94 mm; range, 3.10 - 5.36). In a univariate regression analysis, the LT was associated significantly age, body weight, body height, body mass index (BMI), systolic pressure, diastolic pressure, spherical equivalent (SE), intraocular pressure (IOP), anterior chamber depth (ACD) and axial length (AL) (all *P*<0.05). After adjusting the general parameters and ocular parameters, LT was associated significantly with age (β, 0.0151; 95% CI, 0.0116 - 0.0186; *P*<0.001), gender (β, 0.1233; 95% CI, 0.0553 - 0.1913; *P*<0.001) and ACD (β, -0.5815; 95% CI, -0.8059 - -0.3571; *P*<0.001) using the multivariate regression model. The LT was associated with older age, female gender, and shallower ACD in healthy subjects from Southern China. The data may help understand ocular diseases concerning lens thickness.

## 1. Introduction

The normal crystalline lens is a transparent, biconvex structure, which lies posterior to the iris and anterior to the vitreous body in the globe, playing an essential role in maintaining normal visual function. Previous studies demonstrated that the crystalline lens contributes to approximately one-third of ocular refractive power and takes part in accommodation[1, 2]. In accommodation, the lens thickness (LT) increases by 0.17 to 0.4 mm[3, 4]. Furthermore, the LT still continuously increases by 0.15 to 0.20mm per decade, even though axial length no longer increases[5]. And the raised LT leads to the shallower anterior chamber depth, which would induce the development of angle-closure glaucoma[6]. Therefore evaluation of the LT was helpful to understand the development of some ocular diseases. Though there are many studies about the ocular biometrics, few studies were focused on the LT. Jonas et al. reported the LT and associated factors, yet the subjects in their study were more than 30 years old [7]. To explore the LT of adults and children, we performed the present study to evaluate the LT and associated factors in healthy subjects of southern China.

## 2. Methods

### 2.1 Study design

This was a prospective, observational, single-center, cross-sectional study conducted at the Department of Ophthalmology, Foshan Second Hospital, Southern Medical University, between May 2019 and April 2020. This study was registered on the Chinese Clinical Trial Registry (identifier: ChiCTR1900024921) and approved by the Institutional Review Board of Foshan Second Hospital. The study was in accordance with the tenets of the Declaration of Helsinki. All subjects gave informed consent before the beginning of the study.

### 2.2 Study subjects

Healthy subjects with best-corrected visual acuity (BCVA) of 20/25 or better were included in the study. The inclusion criteria were as follows: age ≥ 5 years, intraocular pressure (IOP) <21 mmHg, normal findings of the slit-lamp examination, and fundus evaluation. The exclusion criteria were: any history of ocular surgery or ocular trauma, opacity media preventing high-quality imaging, glaucoma, systemic diseases, such as hypertension, diabetes mellitus, retinal and choroidal disease, for example, retinal vein obstruction, macular hole, epiretinal membrane, and polychoroidal retinopathy.

### 2.3 Ophthalmic examinations

All subjects underwent a comprehensive ocular examination, including BCVA, slit lamp, IOP, and fundus evaluation. Refraction data were collected with an autorefractor (KR8900, Topcon, Japan). Spherical equivalent (SE; sphere power+cylinder power/2) was used for analysis. IOP was measured three times with noncontact tonometer (TX-20; Canon, Japan) and calculated the mean. Ocular biometric parameters, including central corneal thickness (CCT), axial length (AL), anterior chamber depth (ACD), and LT, were measured with optical low-coherence reflectometry (LenStar LS900, Haag-Streit, Inc., Koeniz, Switzerland). The measurement, which consists of 16 consecutive scans each time, was repeated three times for each eye of all subjects, and the mean was taken for further statistical analysis. The central macular thickness (CMT, central 1 mm) was assessed by the macular cube 512 × 128 model of optical coherence tomography (Cirrus 5000 HD-OCT; Carl Zeiss Meditec, Inc, Dublin, CA). Arterial blood pressure, body weight, and height were recorded, and the body mass index (BMI) was calculated as a variable to explore the association with LT.

### 2.4 Statistical analysis

Data were presented as mean ± standard deviation (SD) for continuous variables and frequency (%) for categorical variables. For the associated factors of LT, regression analysis was performed using a generalized estimation equation (GEE) model that was applied to adjust for the possible intra-eyes correlation. Firstly, univariate GEE was performed to identify potential factors. The dependent variables, including systemic parameters and ocular parameters, were selected into the models, respectively. Then, variables with *P*<0.2 in univariable analysis and potential confounders were included in the multivariate GEE model. Regression coefficients and odds ratios with 95% confidence intervals (CI) were reported. SAS statistical software version 9.4 (SAS Institute Inc., Cary, NC) was used for statistical analysis. *P* value<0.05 was considered statistically significant.

## 3. Results

Of 315 screened individuals, a total of 263 subjects (83.49 %) enrolled. Fifty-two subjects were excluded from analysis, including 12 subjects for hypertension or diabetics, 5 subjects for high ocular pressure, 25 subjects for remarkable cataract, 4 subjects for ocular surgery history, 3 for lack of refraction or biometric data. Finally, the 526 eyes of the 263 remaining subjects were included in the study. The demographic data and clinical characteristics of the subjects and their eyes are summarized in Table 1. The mean LT was 4.01 ± 0.57 mm(95% CI, 3.97 - 4.06; median, 3.94 mm; range, 3.10 - 5.36). The mean age of subjects was 38.48 ± 22.04 years (range, 5 - 84 years). Of the subjects, 138 (52.47%) were female.

In univariate analysis, the LT increased significantly with age (*P*< 0.001, Figure 1), higher body weight (*P*< 0.001), higher body height (*P*< 0.001), higher BMI (*P*< 0.001), lower IOP (*P*=0.02), taller SE (*P*=0.01), shallower ACD (*P*< 0.001, Figure 2), shorter AL (P< 0.001), higher systolic pressure (*P*< 0.001) and higher diastolic pressure (*P*< 0.001). In univariate analysis, LT was not significantly associated with gender (*P*=0.64), CCT (*P*=0.6172), and CMT (*P*=0.41) (Table 2). Variables with P<0.2 in univariable analysis and potential confounders (gender) related to the primary variable included in multivariate models. After controlling the ocular parameters and systemic factors, LT was significantly associated with older age (P< 0.001), female gender (P< 0.001), and shallower ACD (P< 0.001) (Table 3). However, body weight, body height, BMI, SE, AL, systolic pressure, and diastolic pressure were no longer significantly associated with thickness.

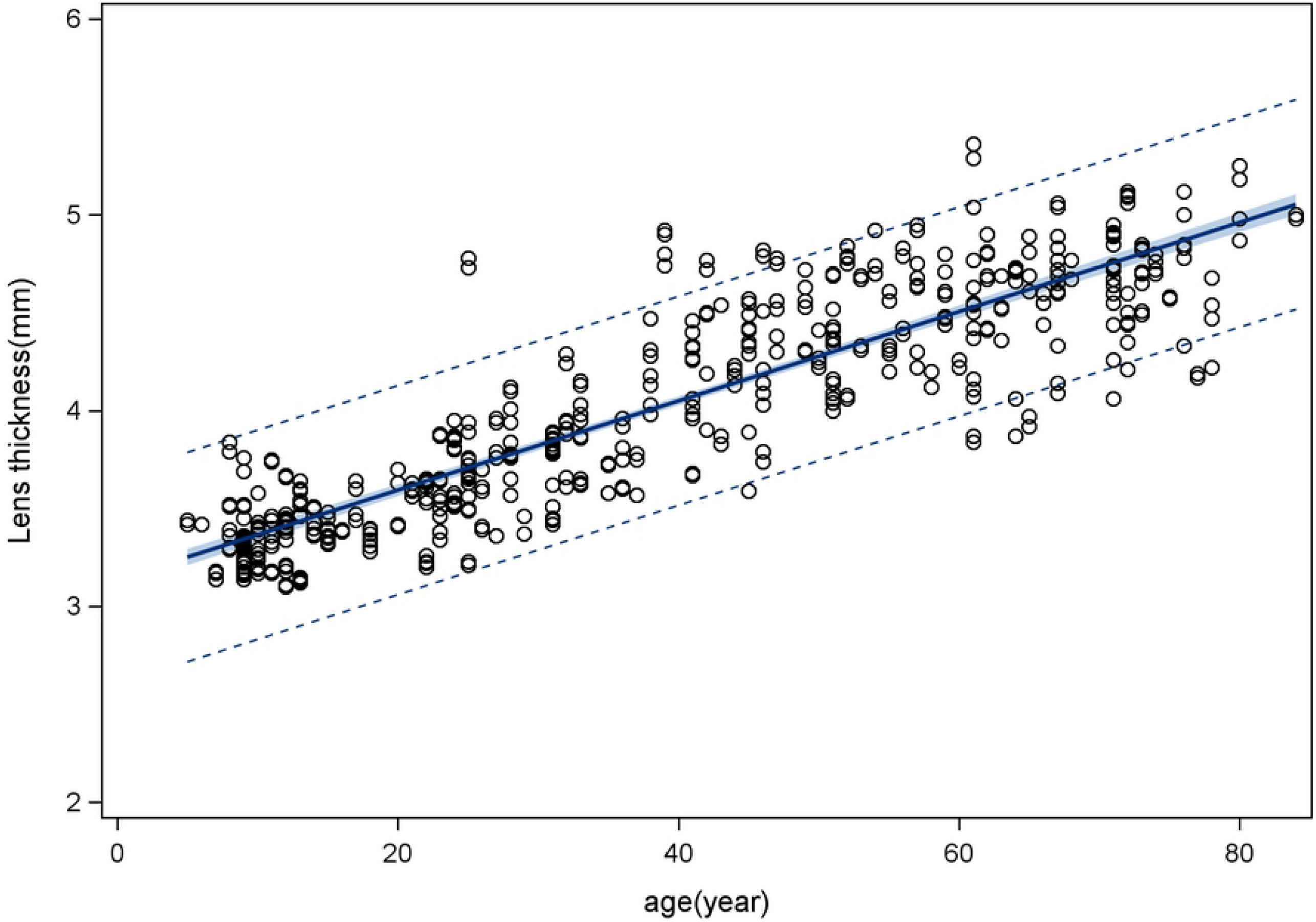

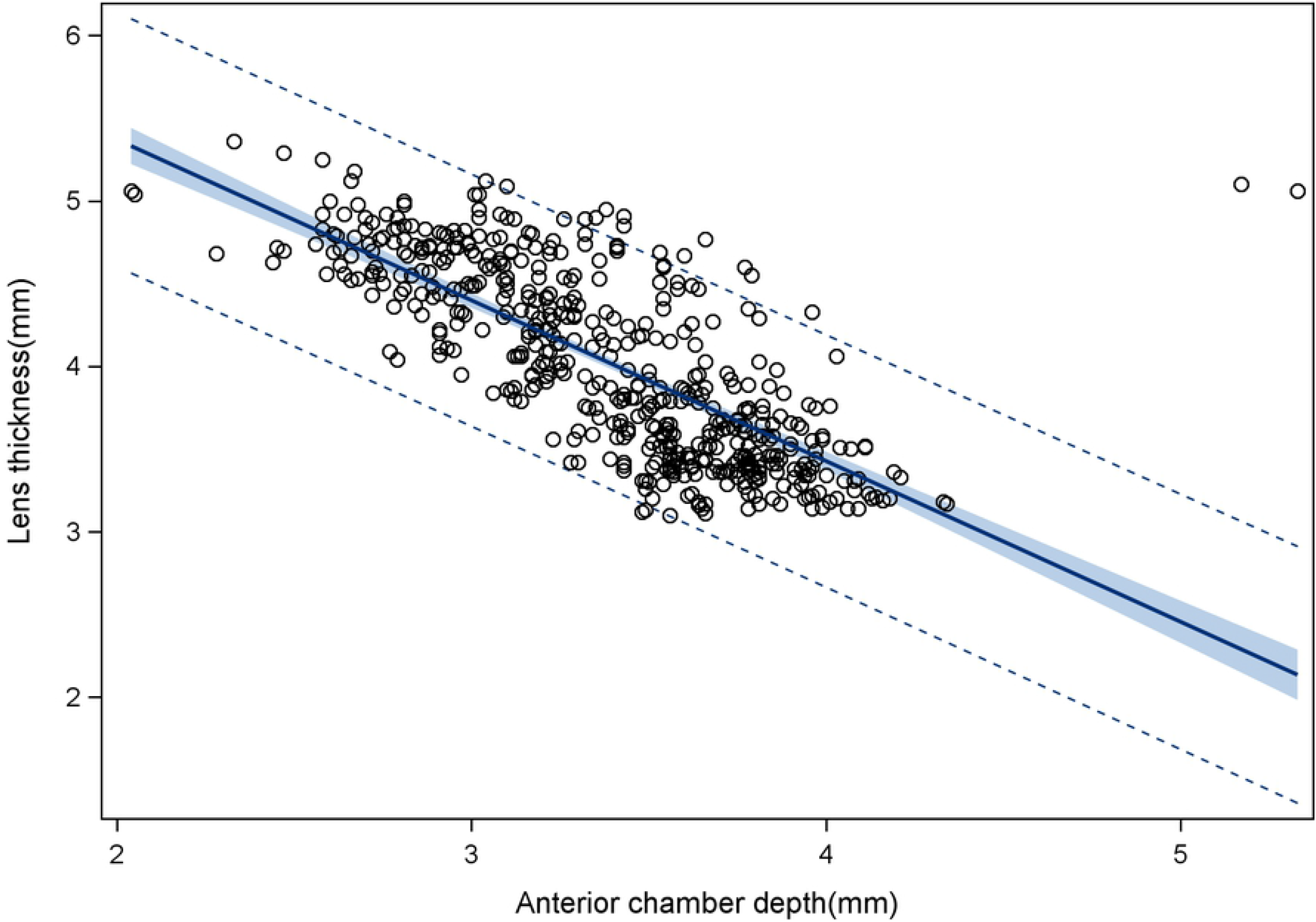

## 4. Discussion

LT, as an essential parameter of the lens, varies from the population and has significance in some ocular diseases, for example, acute angle-closure glaucoma, cataract, and presbyopia[8-10]. However, fewer researches were focused on the LT of healthy subjects. The present study systematically evaluated the LT in healthy Chinese subjects and investigated its association with general and ocular factors. The results show that the mean crystalline LT is 4.01 ± 0.57 mm (95% CI, 3.97 - 4.06; median, 3.94 mm; range, 3.10 - 5.36). It is strongly associated with older age (β, 0.0151; 95%CI, 0.0116-0.0186; *P*< 0.001), female gender (β, 0.1233; 95%CI, 0.0553-0.1913; *P*< 0.001), and shallower ACD (β, -0.5815; 95%CI, -0.8058 - -0.357; *P*< 0.001) after adjusting systemic parameters and other ocular parameters.

The results indicate that healthy subjects from Southern China have similar LT as studies reported previously, although it is a clinic-based cross-sectional study. The mean LT is 4.01 ± 0.57 mm in our study,which is slightly less than the value of 4.07 ± 0.40 mm in the study reported by Lesley Doyle[11] and 4.05 ± 0.20 mm in a study published by Richdale[10]. It is possible relative to the age amplitude. The age range of enrolled subjects is much broader in our research that includes children, and the age of subjects in the previous study was more than 30 years.

It has been proven that the LT was increasing with age based on previous studies. A population-based cross-sectional survey reported by Jonas et al. showed that the mean LT was significantly related to higher age[7] (β=0.003, P<0.001), including 9046 eyes of 4610 subjects with an average age of 49.1 ± 13.2 years (range: 30-100 years). With a per year increase in age, Jonas also reported that the LT maybe increases approximately by 0.009 mm in another study[12]. Similar results were confirmed in other studies[13, 14]. Our results also demonstrated an age-related increase in LT each year (β, 0.0151, *P*< 0.001). Why does the LT increase with age? One explanation was due to the continuous development of the new lens fibers in the equatorial region of the lens[15, 16]. Additionally, the ciliary body becomes relaxed to the curvature enlargement of the lens with aging, which will also cause an increase in LT[17-19].

There was still no consensus with aspect to the association between gender and LT. Jonas’ results showed that the LT was significantly associated with the male gender in a multivariate analysis[7]. In contrast, Hassan et al. reported that LT was positive with the female gender[20]. In our study, LT is predominant in females. The result might explain why angle-closure glaucoma is more common in females than males, especially in older females.

In our study, we found that the ACD would become shallow by 0.58 mm with per 1 mm increase of LT, after adjusting for other variables. The association between the ACD and the crystalline LT was investigated in some studies[15, 21]. Jonas et al. demonstrated that the ACD decreased by 0.007 mm when LT increased by 0.009 mm[12]. In agreement, Praveen et al. also reported that LT would significantly affect the ACD[22]. Still, the Singapore Chinese Eye study demonstrated that LT was not associated independently with ACD[23]. In that study, they thought the lens vault would influence the ACD as well as.

In the present study, we did found the systemic variables, such as height, weight, and BMI were not associated with the LT in the multivariate analysis, although those were associated significantly with LT in univariate analysis. Conversely, in a population-based study with age more than 30 years, authors found that a thicker lens was related to high body stature and high BMI[7], and they considered that tall subjects maybe have larger eyes[24].

The major strength of our study was a relatively large sample size with a wider age range (from 5 to 84 years). Furthermore, we adjusted for the intra-eyes correlation effect on the LT using the GEE model. The limitation of this study should be mentioned. Firstly, it was not a population-based study. The subjects in the study were mainly from ophthalmic clinical, physical examination center, and community screening, which would lead to selection bias to some extent. Secondly, there was no data on LT with an age of less than five years because these children can not finish the examination of ocular biometrics.

In conclusion, the mean LT was 4.01 ± 0.5 mm(95% CI, 3.97 - 4.06 mm)in healthy subjects of Southern China. It was significantly associated with older age, female gender, and shallower ACD after adjusting general and ocular parameters. The results may be helpful to explain the development of ocular diseases as for lens, especially such as cataract and closure-angle glaucoma.

## Data Availability

All relevant data are within the manuscript and its Supporting Information files

## Abbreviations

ACD: anterior chamber depth
AL: axial length
BCVA: best-corrected visual acuity
BMI: body mass index
CCT: central corneal thickness
CI: confidence intervals
CMT: central macular thickness
GEE: generalized estimation equation
IOP: intraocular pressure
LT: lens thickness
SD: standard deviation
SE: spherical equivalent.

## Acknowledgments

We want to express our gratitude to all data collectors and study subjects.

## Auther contributions

Y. Cao and X. Kong conceptualized the study.

Y. Cao performed data analysis and visualization, and wrote the manuscript.

F. Liao performed the procject admistration and data curation.

Y. He collected the data.

Y.Huang provided technical assistance.

L.Zhou revised the manuscript.

All authors read and approved the final manuscript.

### Data Availability

The data of the present study are available from the corresponding author and can be obtained on a reasonable request.

### Conflicts of Interest

The authors have no financial or conflicts of interest concerning this study.

### Funding Statement

This study was no supporte.

## Notes

### Competing Interest Statement

The authors have declared no competing interest.

### Funding Statement

The author(s) received no specific funding for this work.

### Author Declarations

This study was registered on the Chinese Clinical Trial Registry (identifier: ChiCTR1900024921) and approved by the Institutional Review Board of Foshan Second Hospital.

